# Developing a male-specific counselling curriculum for HIV treatment in Malawi

**DOI:** 10.1101/2023.08.08.23293583

**Authors:** Misheck Mphande, Isabella Robson, Julie Hubbard, Elijah Chikuse, Eric Lungu, Khumbo Phiri, Morna Cornell, Sam Phiri, Thomas J Coates, Kathryn Dovel

## Abstract

Men living with HIV in sub-Saharan Africa have sub-optimal engagement in antiretroviral therapy (ART) Programs. Generic ART counselling curriculum in Malawi does not meet men’s needs and should be tailored to men.

We developed a male-specific ART counselling curriculum, adapted from the Malawi Ministry of Health (MOH) curriculum based on literature review of men’s needs and motivations for treatment. The curriculum was piloted through group counselling with men in 6 communities in Malawi, with focus group discussion (FGD) conducted immediately afterward (n=85 men) to assess knowledge of ART, motivators and barriers to care, and perceptions of the male-specific curriculum. Data were analysed in Atlas.ti using grounded theory. We conducted a half-day meeting with MOH and partner stakeholders to finalize the curriculum (n=5).

The male-specific curriculum adapted three existing topics from generic counselling curriculum (status disclosure, treatment as prevention, and ART side effects) and added four new topics (how treatment contributes to men’s goals, feeling healthy on treatment, navigating health systems, and self-compassion for the cyclical nature of lifelong treatment. Key motivators for men were embedded throughout the curriculum and included: family wellbeing, having additional children, financially stability, and earning/keeping respect. During the pilot, men reported having little understanding of how ART contributed to their personal goals prior to the male-specific counselling. Men were most interested in additional information about treatment as prevention, benefits of disclosure/social support beyond their sexual partner, how to navigate health systems, and side effects with new regimens. Respondents stated that the male-specific counselling challenged the idea that men were incapable of overcoming treatment barriers and lifelong medication.

Male-specific ART counselling curriculum is needed to address men’s specific needs. In Malawi context, topics should include: how treatment contributes to men’s goals, navigating health systems, self-compassion/patience for lifelong treatment, and taking treatment while healthy.

## Introduction

Men in sub-Saharan Africa are less likely than women to start and stay on HIV treatment, and as a result have higher morbidity, mortality and rates of viremia as compared to women [1–3]. Men also have lower knowledge about the benefits of starting antiretroviral therapy (ART) early and engaging in sustained care over one’s life time, and how to manage treatment side effects, fear of unwanted disclosure, and other barriers to sustained treatment engagement [4, 5]. There are few interventions that specifically address men’s unique needs related to sustained ART engagement [6, 7].

Standard ART-related counselling in the region is not tailored to sub-population needs, including men. Unintentionally, dyadic and information focused forms of counselling can reinforce fears about lifelong treatment through messages that focus on what clients must give up to be on treatment (such as alcohol, multiple partners, and adhering to specific diets and having to keep strict schedules for taking medication). [8]. ART-related educational materials are often biomedically focused and do not address the social and behavioral benefits and support strategies for lifelong treatment [9]. Patient-centered, responsive counselling is needed that draws on core components of motivational interviewing, whereby counsellors help collaborate with individuals to identify and strengthen their personal motivations for change [10].

Men face unique barriers to sustained treatment engagement that are often not addressed in standard counselling, resulting in limited motivation to remain in care. These barriers include fear of ART side effects, perceived stigma and unwanted disclosure as well as work commitments that conflict with ART clinic hours and threaten their ability to generate income [11]. Many men experience general and didactic ART counselling as demoralizing and judgmental, and describe language used during counselling sessions as overly technical and paternalistic [12, 13]. Purely didactic counselling can also inhibit open enhance between client and HCW, discouraging clients from expressing their concerns or brainstorming personalized solutions to challenges [14] hence the need for motivational counselling. A growing number of studies show that male-specific ART counselling that is responsive to men’s needs and priorities can improve ART retention [15, 16]. Yet, there is little guidance on how to modify existing counselling curricula for targeted specific populations such as men.

We developed and implemented male-specific ART counselling and education materials for men living with HIV (MLHIV) in Malawi. The curriculum targeted men who either never initiated ART or who initiated but had experienced treatment interruption. In this paper we describe the steps taken to develop a male-specific curriculum, modifying the existing Ministry of Health (MoH) curriculum, and the final components of male-specific counselling tailored to men’s needs in Malawi.

## Methods

We adapted the Centers for Disease Control and Prevention’s (CDC) guidance on the adaptation process for evidence–based behavioral interventions (EBI) within the field of HIV [17]. The CDC guidelines propose that curriculum should be adapted in five steps, which we modified to fit the local context (see Table 1).

**Table 1:** Step by Step development of male-specific counselling curriculum.

| **CDC Guidance on Adaptation of EBI** | **Steps Completed by Our Team** |
| --- | --- |
| 1. Assess needs of the population | 1. Scoping review on specific needs of men for ART engagement |
| 2. Select the intervention to be modified | 2. Review MoH ART counselling materials against specific needs of men from step 1 |
| 3. Make adaptations needed based on the initial | 3. Adapt MoH counselling materials based on steps 1 and 2 |
| 4. Pilot the adapted intervention | 4. Pilot the adapted curriculum with men<br>5. Review the finalized curriculum with stakeholders and implementers |
| 5. Implement the adapted intervention | 6. Implement the final adapted curriculum |

Below we describe each step taken to modify the MoH curriculum into a male-specific curriculum (Table 1).

### Step 1. Scoping review on specific needs of men for ART engagement

We first conducted a scoping review of published, grey and white literature from southern and eastern Africa to identify facilitators and barriers to men’s engagement in the HIV cascade, prioritizing findings from Malawi. We conducted a search on Google Scholar and reviewed documents on men’s HIV care from stakeholders including UNAIDS, WHO and PEPFAR.[18, 19] The process was guided by three primary questions: what are male-specific 1) barriers, 2) facilitators, and 3) knowledge gaps affecting ART (re-) initiation and retention?

### Step 2. Review MoH ART counselling materials against specific needs of men from step 1

We used findings from step 1 to systematically identify gaps and recommendations to existing ART counselling curriculum. We reviewed existing MoH ART counselling curriculum to identify areas where the specific needs of MLHIV identified in the scoping review were not adequately addressed. First, the lead author (MM) entered MoH counselling content by topic into an Excel data capture tool. For each topic, we entered data on the following categories: 1) What was the main message; 2) How did the content (subject, wording, structure, graphic) meet (or miss) male-specific needs; 3) What evidence supports this assessment from the scoping review; and 4) if gaps were identified, what (if any) content is supported by evidence in the scoping review (i.e. specific wording, messaging, graphics, etc.). Co-authors (KD, JH, IR) independently reviewed and gave feedback to the data capture tool until we reached agreement on all categories.

### Step 3. Adapt MoH counselling materials based on steps 1 and 2

We adapted the MOH curriculum based on the finalized data extraction and assessment conducted in Step 2. Local Malawian team members developed detailed changes and/or additions to curriculum and entered suggested adaptations into the same Excel data capture tool. All initial adaptations were reviewed by co-authors and underwent multiple iterations until consensus was reached. We used 4 in-person group meetings to promote discussion and consensus building.

Upon consensus, the adapted curriculum was extracted from the data capture tool and formatted into a small, flip-chart style job aid for health care worker (HCW) and client use during counselling sessions, following standard formatting for counselling curriculum in Malawi. The draft male-specific counselling curriculum was translated into the local language and male-specific graphics were created by a local, graphic designer.

### Step 4. Pilot adapted curriculum with men

We piloted the adapted draft male-specific counselling curriculum with 86 men with unknown HIV status. We recruited men through convenience sampling from six villages in central (n=3 villages) and southern (n=3 villages) regions of Malawi. Men were eligible to participate if they 1) resided in the village and 2) were ≥15 years old. The counselling curriculum was presented in a group counselling format by two trained Research Assistants and followed immediately by Focus Group Discussions (FGD). FGDs assessed men’s post-counselling knowledge of ART, motivators and barriers to care, and perceptions of the adapted male-specific curriculum. Participants were asked if the adapted curriculum was relevant to their lives and their motivations as men. They were asked to identify any topics that were unclear or not motivating in the curriculum, and additional needs or questions they still had about HIV services. They were also invited to suggest any further modifications needed, including specific language and graphic choices to fit the local context. FGDs were conducted in the local language (Chichewa), translated to English, transcribed, and analyzed in Atlas.ti8 [20] using grounded theory [21]. Participant responses were reviewed, main themes summarized, and the curriculum further modified to incorporate feedback. Verbal informed consent was obtained prior to the interviews.

### Step 5. Review of finalized curriculum

The male-specific ART counselling curriculum was reviewed by two groups: programmatic stakeholders in Malawi and HWCs who would implement the new curriculum.

#### Stakeholders

In a one-day stakeholder workshop, the updated male-specific counselling curriculum flip chart and artwork was presented to PEPFAR implementing partner staff (n=5) and HIV researchers (n=6). Participants reviewed the content (including topics, analogies, wording, and graphics) to ensure the curriculum complied with MOH guidelines for HIV counselling, and assess the curriculum’s feasibility and acceptability among stakeholders. The curriculum was then modified based on consensus.

#### Implementers

Finally, male nurses (n=10) and a lay cadre (treatment supporters, n=10) were trained in the new curriculum and invited to provide further modifications to improve clarity and implementation. The training took place over a five-day period and all modifications were agreed upon by consensus. Final edits were made prior to implementing the male-specific counselling within the parent trials.

### Step 6. Implement the final adapted male-specific counselling curriculum

The final male-specific curriculum was implemented as part of two randomized control trials (IDEAL and ENGAGE). Results of the implementation will be described in in the trials’ primary outcome papers.

## Results

### Step 1: Scoping review on specific needs of men for ART engagement

We identified eight overarching male-specific barriers and facilitators affecting men’s ART re-initiation and retention (Table 2). First, men desired to be engaged as equals with HCWs, valuing respect and autonomy over care[11, 22]. Men did not respond well to shouting or conflictual interactions with HCWs, or counselling sessions in which they are simply given instructions without being given motivational explanations or choice [23–25] Men desired interactive counselling sessions, and to be treated as equal participants in their own health care [25]. Men also needed to have a sense of ownership and agency over their own health [16]. Without positive interactions with HCWs, men often opted to avoid services altogether in order to maintain respect and autonomy [22].

**Table 2:** Barriers and facilitators affecting men’s ART (re-)initiation and retention on ART in Malawi.

| No | Theme | Barrier/Facilitator | Description |
| --- | --- | --- | --- |
| 1 | Masculinity | Barrier | Expectations, norms and roles influence men's decisions about starting treatment, disclosing their status and remaining in care |
| 2 | Men as economic Pillars/ competing interests | Barrier | Men see themselves as economic pillars, they want to see the value of ART in their income- generating activities |
| 3 | Limited ART knowledge/ Side effects | Barrier | Knowledge gaps on TASP, side effects and flexibility about taking ART |
| 4 | Good health | Facilitator | Men want to have good health to maintain their strong physical appearance and reputations in the community |
| 5 | Gendered service provision/ preferred gender matching service provider | Barrier/Facilitator | Men are less familiar with health systems / Men would use the services if provided by HCWs of their own gender |
| 6 | Autonomy | Facilitator | Men want to be engaged in and have ownership over their HIV care |
| 7 | Social support/ guardian support | Facilitator | Men perform better on treatment if they have a role model, and support, especially from a fellow man |
| 8 | Self-compassion | Facilitator | Clients who have self-compassion are likely to have better adjustment to treatment, less stress, positive health seeking attitudes, and are more likely to disclose status and practice safe-sex |

Second, extensive literature framed men as the economic pillar of the household [26, 27]. Bearing the responsibility for income generation was both a barrier and facilitator for men. Income generation acted as a barrier because men may interrupt treatment due to travel for work[5, 28, 29] and conflicts between clinic and work schedules[11, 30]. Failing to provide for their families was also interpreted as weakness and directly undermines men’s internalized expectations of masculinity[27]. However, income generation facilitated men using ART services, as men cared about providing for their families could clearly see how ART contributed to their earning potential[31–33].

Third, men were motivated by maintaining their own health [31, 34]. Health was important from several perspectives: good health allowed men to keep their HIV status private from community members, maintaining their reputation in the community. Good health is also associated with a strong physical appearance which increases men’s options for sexual partners[27, 35]. Finally, physical strength and vitality are seen as directly influencing men’s earning potential, key to their current and future role as provider for their family [2, 7, 35, 36].

Fourth, the gendered organization of HIV services means that men are less familiar with health facility protocols and strategies to overcome facility-level barriers to care [Hubbard 2022]. Men often reported difficulties navigating health facilities, especially if the majority of services provided at facilities is designated for women and children[7, 16, 37, 38].Many men often preferred to be offered HIV services by fellow male HCWs, and gender-sensitive services can facilitate service utilization for men [27].

Fifth, men had limited knowledge about ART services that could increase utilization [5, 36, 39]. Men were still largely unaware of TasP strategies and the benefits of early ART initiation[5], and had limited comprehension about drug doses and how HIV and ART work within the body. Men over-estimated the risk of severe side effects when taking ART, leading to concerns that ART would undermine their health. Men were also concerned that such side effects might impact their physique, a key barrier for men’s slow uptake of ART services [40, 41]. While side effects are rarely experienced with new dolutegravir-based regimens [42, 43], the legacy of side effects due to old regimens (such as nausea, bad dreams, and fat deposits) still informs men’s fears of treatment today [44].

Sixth, social support from family members or male friends was critical to men’s sustained retention in ART services. Fear of disclosure of one’s status and the subsequent anticipated stigma is a common reason for defaulting from ART[11]. Recent studies suggest that having a male role model who has successfully engaged in ART and is thriving in care may be particularly motivating for men[25, 30, 45].

Seventh, literature suggested that self-compassion with life-long treatment results in better adjustment and lower stress, anxiety and shame[32, 46]. Self-compassion facilitates HIV status disclosure, encouraging clients to practice safe sex and seek medical care [46]. We defined self-compassion as being able to acknowledge the struggles one faces and celebrate the small gains achieved along the journey. However, men may have limited self-compassion as harmful gender norms define men as being strong, self-sufficient and never weak[27, 32]. Self-compassion with lifetime treatment may help alleviate fears that prevent some men from ever starting treatment and may remove the feeling of being overwhelmed by treatment.

Finally, our findings strongly supported the literature that masculinities play a huge role in men’s engagement in treatment and need to be explored [5, 27, 47]. A meta-analysis showed that beliefs about masculinity may influence treatment outcomes [48]. For example, in South Africa, men considered being tough, unemotional, aggressive, denying weakness, sexually unstoppable and appearing physically strong as important male traits [27, 32]. These masculinity constructs are likely to prevent some men seeking health care services [26, 32, 39].

### Step 2: Review MoH counselling materials for gaps in reaching men

We found that the MoH ART counselling curriculum was didactic and directive. The curriculum was largely biomedical-focused, explaining what ART does and the need for adherence, but not explicitly linking how ART can contribute to individuals’ goals and needs (see Table 3 for the completed data capture tool). The curriculum did not provide explicit probes needed to facilitate an interactive or client-centered counselling.

**Table 3:**
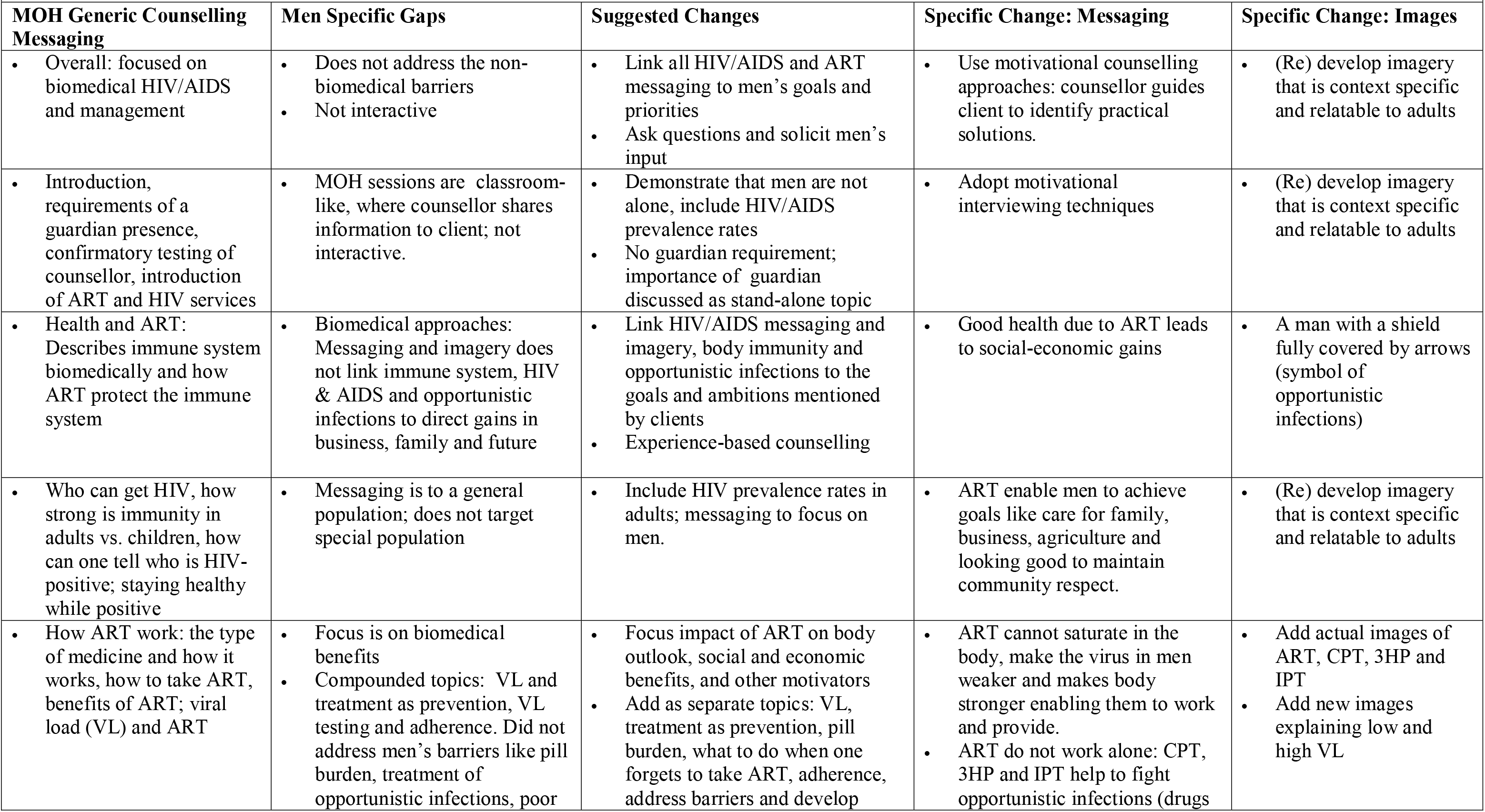
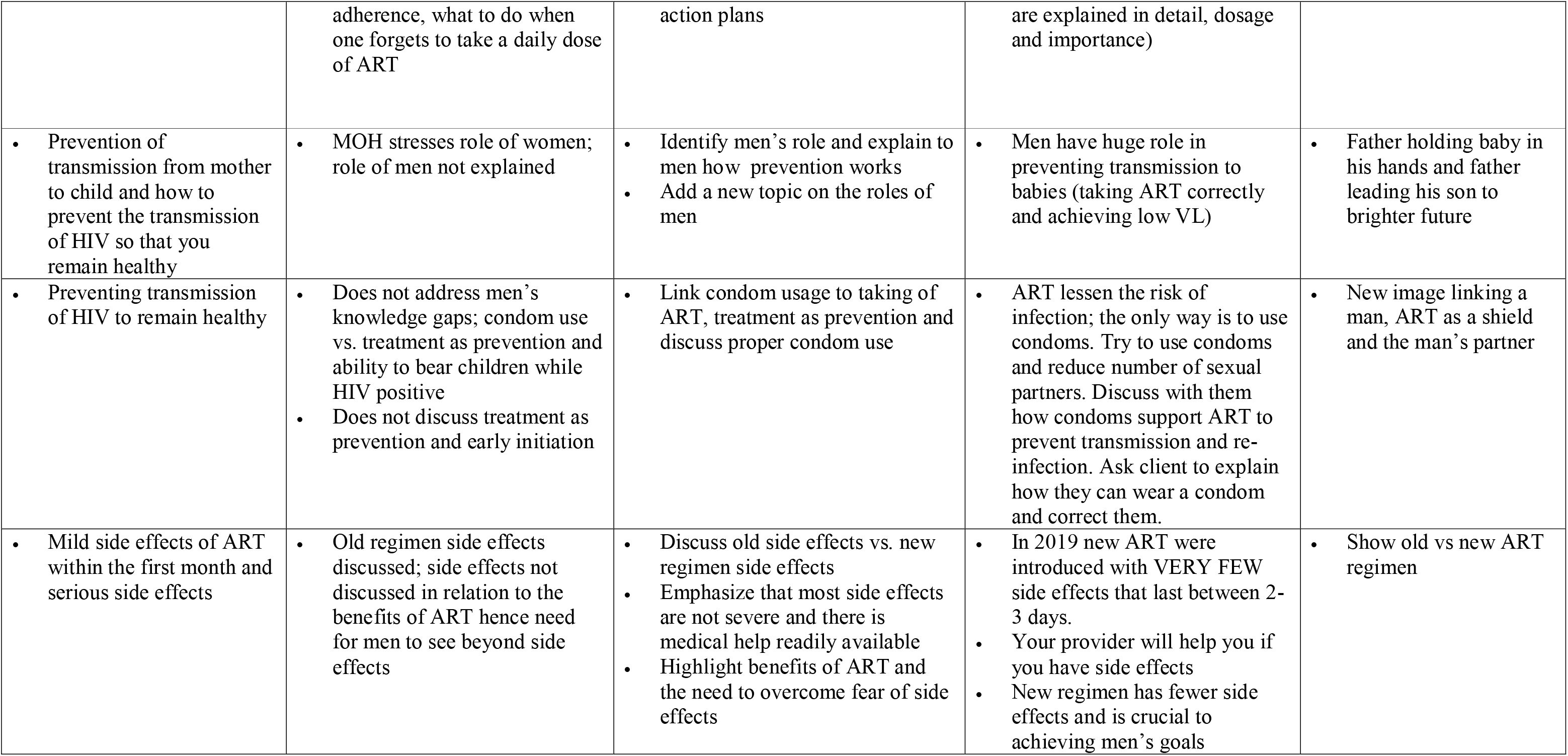
Modified Topics in the adapted counselling pamphlet.

| **Table 3: Modified Topics in the adapted counselling pamphlet** |  |  |  |  |
| --- | --- | --- | --- | --- |
| **MOH Generic Counselling Messaging** | **Men Specific Gaps** | **Suggested Changes** | **Specific Change: Messaging** | **Specific Change: Images** |
| Overall: focused on biomedical HIV/AIDS and management | Does not address the non-biomedical barriers<br>Not interactive | Link all HIV/AIDS and ART messaging to men's goals and priorities<br>Ask questions and solicit men's input | Use motivational counselling approaches: counsellor guides client to identify practical solutions. | (Re) develop imagery that is context specific and relatable to adults |
| Introduction, requirements of a guardian presence, confirmatory testing of counsellor, introduction of ART and HIV services | MOH sessions are classroom-like, where counsellor shares information to client; not interactive. | Demonstrate that men are not alone, include HIV/AIDS prevalence rates<br>No guardian requirement; importance of guardian discussed as stand-alone topic | Adopt motivational interviewing techniques | (Re) develop imagery that is context specific and relatable to adults |
| Health and ART: Describes immune system biomedically and how ART protect the immune system | Biomedical approaches: Messaging and imagery does not link immune system, HIV & AIDS and opportunistic infections to direct gains in business, family and future | Link HIV/AIDS messaging and imagery, body immunity and opportunistic infections to the goals and ambitions mentioned by clients<br>Experience-based counselling | Good health due to ART leads to social-economic gains | A man with a shield fully covered by arrows (symbol of opportunistic infections) |
| Who can get HIV, how strong is immunity in adults vs. children, how can one tell who is HIV-positive; staying healthy while positive | Messaging is to a general population; does not target special population | Include HIV prevalence rates in adults; messaging to focus on men. | ART enable men to achieve goals like care for family, business, agriculture and looking good to maintain community respect. | (Re) develop imagery that is context specific and relatable to adults |
| How ART work: the type of medicine and how it works, how to take ART, benefits of ART; viral load (VL) and ART | Focus is on biomedical benefits<br>Compounded topics: VL and treatment as prevention, VL testing and adherence. Did not address men's barriers like pill burden, treatment of opportunistic infections, poor | Focus impact of ART on body outlook, social and economic benefits, and other motivators<br>Add as separate topics: VL, treatment as prevention, pill burden, what to do when one forgets to take ART, adherence, address barriers and develop | ART cannot saturate in the body, make the virus in men weaker and makes body stronger enabling them to work and provide.<br>ART do not work alone: CPT, 3HP and IPT help to fight opportunistic infections (drugs | Add actual images of ART, CPT, 3HP and IPT<br>Add new images explaining low and high VL |
|  | adherence, what to do when one forgets to take a daily dose of ART | action plans | are explained in detail, dosage and importance) |  |
| Prevention of transmission from mother to child and how to prevent the transmission of HIV so that you remain healthy | MOH stresses role of women; role of men not explained | Identify men's role and explain to men how prevention works<br>Add a new topic on the roles of men | Men have huge role in preventing transmission to babies (taking ART correctly and achieving low VL) | Father holding baby in his hands and father leading his son to brighter future |
| Preventing transmission of HIV to remain healthy | Does not address men's knowledge gaps; condom use vs. treatment as prevention and ability to bear children while HIV positive<br>Does not discuss treatment as prevention and early initiation | Link condom usage to taking of ART, treatment as prevention and discuss proper condom use | ART lessen the risk of infection; the only way is to use condoms. Try to use condoms and reduce number of sexual partners. Discuss with them how condoms support ART to prevent transmission and re-infection. Ask client to explain how they can wear a condom and correct them. | New image linking a man, ART as a shield and the man's partner |
| Mild side effects of ART within the first month and serious side effects | Old regimen side effects discussed; side effects not discussed in relation to the benefits of ART hence need for men to see beyond side effects | Discuss old side effects vs. new regimen side effects<br>Emphasize that most side effects are not severe and there is medical help readily available<br>Highlight benefits of ART and the need to overcome fear of side effects | In 2019 new ART were introduced with VERY FEW side effects that last between 2-3 days.<br>Your provider will help you if you have side effects<br>New regimen has fewer side effects and is crucial to achieving men's goals | Show old vs new ART regimen |

**Table 3_b:** Newly Introduced Topics in the adapted counselling pamphlet.

| Missing in MOH Generic Counselling Messaging | Men Specific Gaps | Suggested Changes | Specific Change: Messaging | Specific Change: Images |
| --- | --- | --- | --- | --- |
| • Treatment as prevention | • MOH focus is on condom use and reducing number of partners | • Explain how treatment as prevention works | • If you take ART daily, it is unlikely that HIV will be passed to your partner | • (Re) develop imagery that is context specific and relatable to adults |
| • If you forget taking your ART | • MOH is rigid and stresses taking ART same day and same time | • Explain to the client the scientifically allowed time to take ART | • If you miss taking ART, take immediately; if more than 12 hours late, do not take ART, take the next day | • Picture of man worried about having missed taking ART, next to a man taking ART |
| • Experiences with health facilities | • MOH does not address facility level barriers | • Introduce facility client services like health advisory committee; highlight client's rights and responsibilities | • You seek redress from Health Advisory Committee, a different provider or seek transfer out | • Men on waiting area with HAC on the wall |
| • Male guardian support | • MOH stresses on general guardians the importance of having a male guardian; role of male guardian not highlighted | • Discuss role of male guardian and identify who can be the client's male guardian | • With client, identify which male guardian would be central to their treatment | • Different scenarios of a man helping another man |
| • Alcohol | • MOH stresses on stopping alcohol; does not explain why clients must stop and what to do when struggling with alcohol | • Assess client's knowledge and habits; discuss how alcohol might impact efficiency of ART. | • Even if you don't plan to stop taking alcohol, start taking ART | • Scales of balance of taking one bottle of alcohol vs consuming more |
| • Mobility | • MOH barely discusses mobility and ART, emergency refills and adherence | • Discuss emergency refills and the requirements for travelling | • Know your regimen, you can get emergency refills at any facility, you can get a refill before your appointment date | • A man packing for a trip, with ART and health passport book on the items to be packed |
| • Self-compassion and treatment | • MOH does not address challenges in the lifelong/cyclic treatment journey | • Acknowledge decision is difficult and journey has challenges<br>• Establish that there is a way to manage barriers and there is support to slowly but steadily achieve | • Taking ART for life is challenging, but slowly one can learn and overcome challenges | • A picture of a provider helping a client climb a mountain |

The MoH curriculum did not address many of the specific barriers and facilitators experienced by men. There was little consideration of ART in relation to men’s overarching goals and fears. Another gap in the curriculum was the limited discussion on social support. Lastly, the curriculum presented a strict approach to ART adherence, without explaining that missed doses could be taken subsequently and did not include any information about self-compassion while navigating lifelong medication, or any Welcome Back messaging in case individuals fell out of care in the future. Based on this assessment, multiple messaging and graphic changes were suggested

### Step 3: Adapt MoH counselling materials

The draft counselling curriculum adapted to male-specific needs included four new themes and three modified themes (see Table 4). Key messaging included ART as a way for men to regain health, provide for their families, maintain their role within their community, and to have self-compassion with their own ART adherence and facility attendance as life is complicated and lifelong adherence can be difficult.

**Table 4:** Modified and new themes in the male-specific counselling pamphlet.

| Theme | Male-Specific Modification |
| --- | --- |
| New themes |  |
| How treatment contributes to men's goals | We work with men to frame their possible future goals based on their health. Prior to this, men argued that ART engagement competed with business and agriculture priorities. We discuss how HIV services contribute to their key goals. We employ motivational interviewing skills to discussing ART as an integral component to achieve goals. |
| Feeling healthy on treatment and low ART knowledge | We explicitly acknowledge challenges of taking ART when feeling healthy, and ask men to reflect on their own experience. We discuss how taking ART while healthy will prevent disruption of earning prospects and support strong business and better families, using local analogies and graphics that resonate with Malawian men. |
| Navigating health system | We discuss barriers men have experienced when seeking health services at facilities and discuss how to overcome them, and notably, to report poor services. We address the pre-existing discord between HCWs and men. |
| Self-compassion/patience for lifelong treatment | We normalize the fact that men may forget a dose, feel guilty and panic. We highlight the importance of returning to care as soon as possible. We discuss alcohol use and fears about long-term treatment adherence. We acknowledge individual concerns about competing responsibilities, fear of disclosure, and treatment fatigue, and stress that such fears are normal. |
| Modified Themes |  |
| Status disclosure to male friends/family | Beyond disclosure to their sexual partners, we highlight the importance of disclosing to their male social support system. We discuss who is important to them, how to disclose to other men, and how to get social support for ART disclosure. |
| Treatment as prevention | In addition to the general treatment as prevention message, we provide a detailed description about how treatment as prevention works, men's role in preventing vertical transmission, and how these concepts contribute to men's role as provider and building a stronger future. |
| Understanding ART (Side effects) | MOH counselling simply describes potential side effects and tells clients to report to clinic if they persist. We focus on the changes in ART regimen including that the new dolutegravir regimen has fewer side effects than the older regimens |

The counselling curriculum was printed in a small, flip chart style job aid. In developing the job aid, the artist featured a central male character throughout the curriculum. The character was intended to be relatable and a role model figure to most men. He was strong and clean but simply dressed like a wealthy man living in the village, and middle-aged (indicated by a few grey hairs). The male character was depicted in various scenarios that were identified as motivating in the scoping review. For example, being healthy and providing for one’s family were depicted through the image of the main character farming, and looking strong.

### Step 4: Pilot modified male-specific counselling curriculum

In piloting the male-specific counselling curriculum, Men believed that the new curriculum directly connected with what they cared about: their families’ well-being and their role as provider.

> *“It [the curriculum] is connecting to our income generating activities because [it makes it clear] that if we are HIV positive and we adhere to the treatment, we can be strong enough to do our various works and make money in the process and take care of our families.”- FGD*

Men appreciated the fact that the new counselling was interactive and solicited men’s voices, and contrasted this experience with the directive and authoritarian counselling styles to they are accustomed. Men did not like the classroom approach of standard counselling and wished to be fully informed and involved in interactive discussions and decision-making about their health. One man reported:

> *“[with the usual counselling] HCWs came and counselled us without telling us the topic of discussion. They should have told us what we did wrong and later on tell us the negative effects of what we were indulging in.”- FGD*

Men reported numerous curriculum topics that were motivating. Most men were unaware of TaSP and were highly motivated by the fact that taking ART meant that men living with HIV could have HIV-negative children.

> *“A new thing that I have learnt today is that if we are expecting a child and we both are HIV positive, taking the treatment as recommended makes sure that the baby to be born would not get infected. I will even teach others about it.”- FGD.*

Men were also motivated by hearing that the new ART regimen (Dolutegravir) had reduced side effects. Prior to the counselling, many men still associated ART with serious side effects that could inhibit work and income generation, especially if taken while a man was still healthy. Most men believed that the advances of Dolutegravir, including reduced side effects and drug toxicity, needed to be thoroughly communicated with men in the community to improve men’s acceptance of treatment.

> *“We are happy with the messaging on ART [in the new curriculum] because some of us were concerned that when you start taking the ART you have a lot of side effects. But today we learnt that the new drugs have no side effects and one can take them without any problems “-FGD.*

Men were also largely unaware that there was any flexibility about the time when they took ART. Most men narrated strict times of taking daily doses. They were unaware that if they missed a dose, there is an allowance of taking ART within 12 hours. This flexibility was important because it helped men feel like they could reach “good enough” adherence without having to restructure their lives around taking HIV treatment, making adherence attainable for busy men with competing priorities.

> *“The new thing that I have heard is that in the past we had knowledge that people on medication take them only at night, but we have learnt today that they can be taken at any time of the day.”- FGD.*

Men had several suggestions to further refine the curriculum. First, they suggested that local analogies and popular phrases be used throughout the curriculum so that men could easily relate to the topics. For example, in the topic “taking ART while healthy” men suggested using the analogy of a house that is slowly being attacked by termites (the virus in a healthy body), which, if not treated in time will fall completely (the development of the stage of AIDS). For the topic, “Lifelong treatment”, they proposed an analogy of the persistence of a grasshopper which slowly hops (small gains day by day) to cover long distances (see fig A).

**Fig A:**
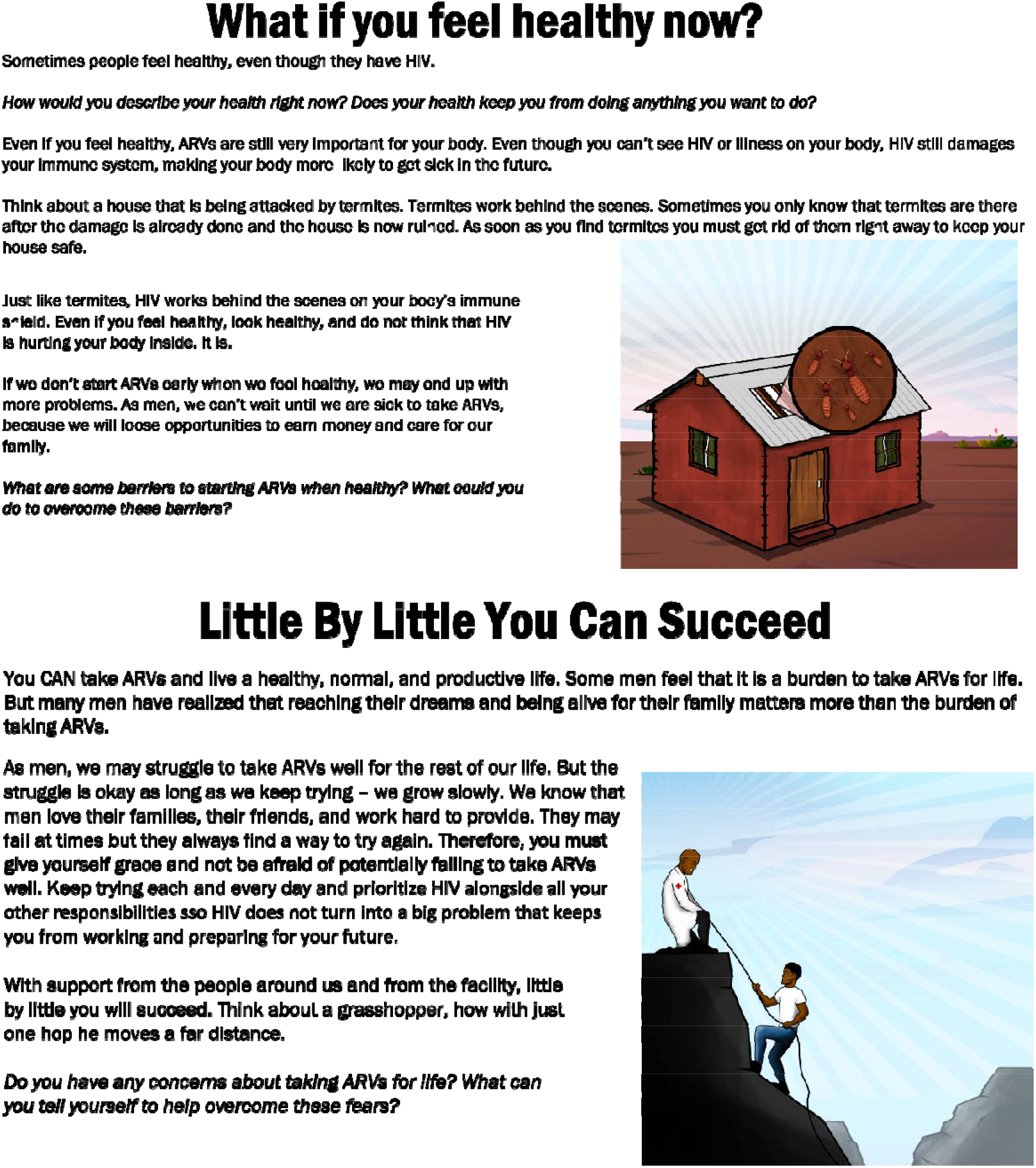
Slides on Taking ART while healthy” and “self-compassion topics

Men also desired graphics throughout the counselling job aid that included men they could relate to (see Fig B).

**Fig B:**
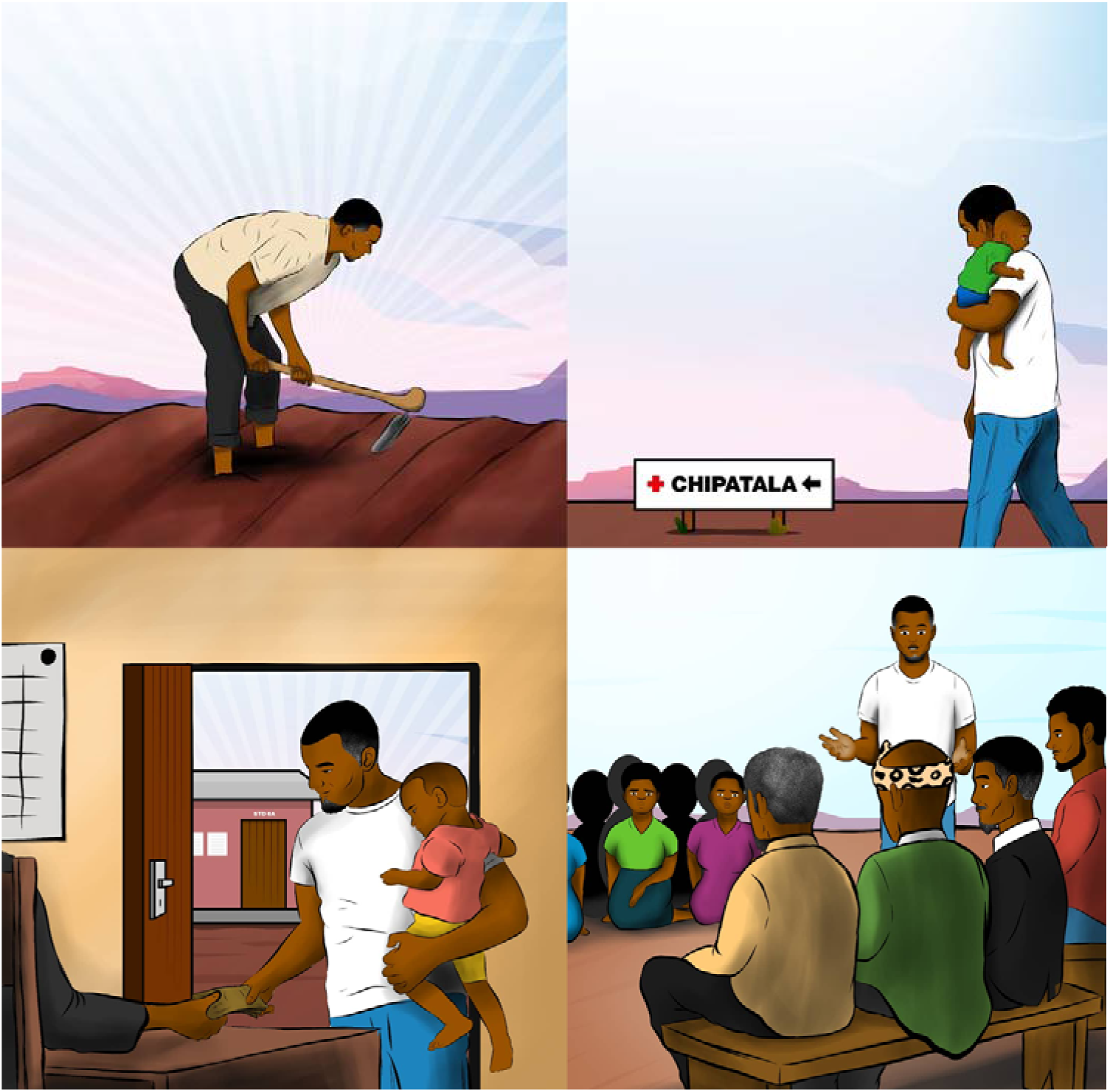
An image showing men as pillars of their society and homes

> *I think the pictures would also help because at school when we are teaching a child we use pictures to illustrate what is going on and how to identify things. Pictures would help people to understand the benefits.”- FGD.*

> *“Give us a picture showing the drugs so that we should be able to show some men can they can see the difference between the old and new medication.”- FGD.*

Based on the FGD’s, the curriculum was further adapted, with emphasis on TasP, reduced side effects of the new ART regimen, ART ability to support men’s overarching goals, and the inclusion of local analogies and graphic visualizations of relatable men.

### Step 5: Stakeholder Review of Finalized Curriculum

Stakeholders and implementers made further small modifications to content and graphics. Both stakeholders and implementers were concerned about the large emphasis on TaSP with fears that it may increase risky sexual behavior. After multiple rounds of discussion, we minimized TaSP messaging and removed language that did not explicitly encourage condom use. A near-final version was reviewed with facility and program staff, nurses and lay-cadre.

### Step 6: Final Product and Implementation Strategies

The final product was a 17-topic counselling pamphlet. The counselling materials included a script for every topic with key bullets to guide HCWs, and probes that promoted client-centered approaches to counselling, including open-ended questions and motivational explanations for how the topic may help men achieve their life goals (see Fig C).

**Fig C:**
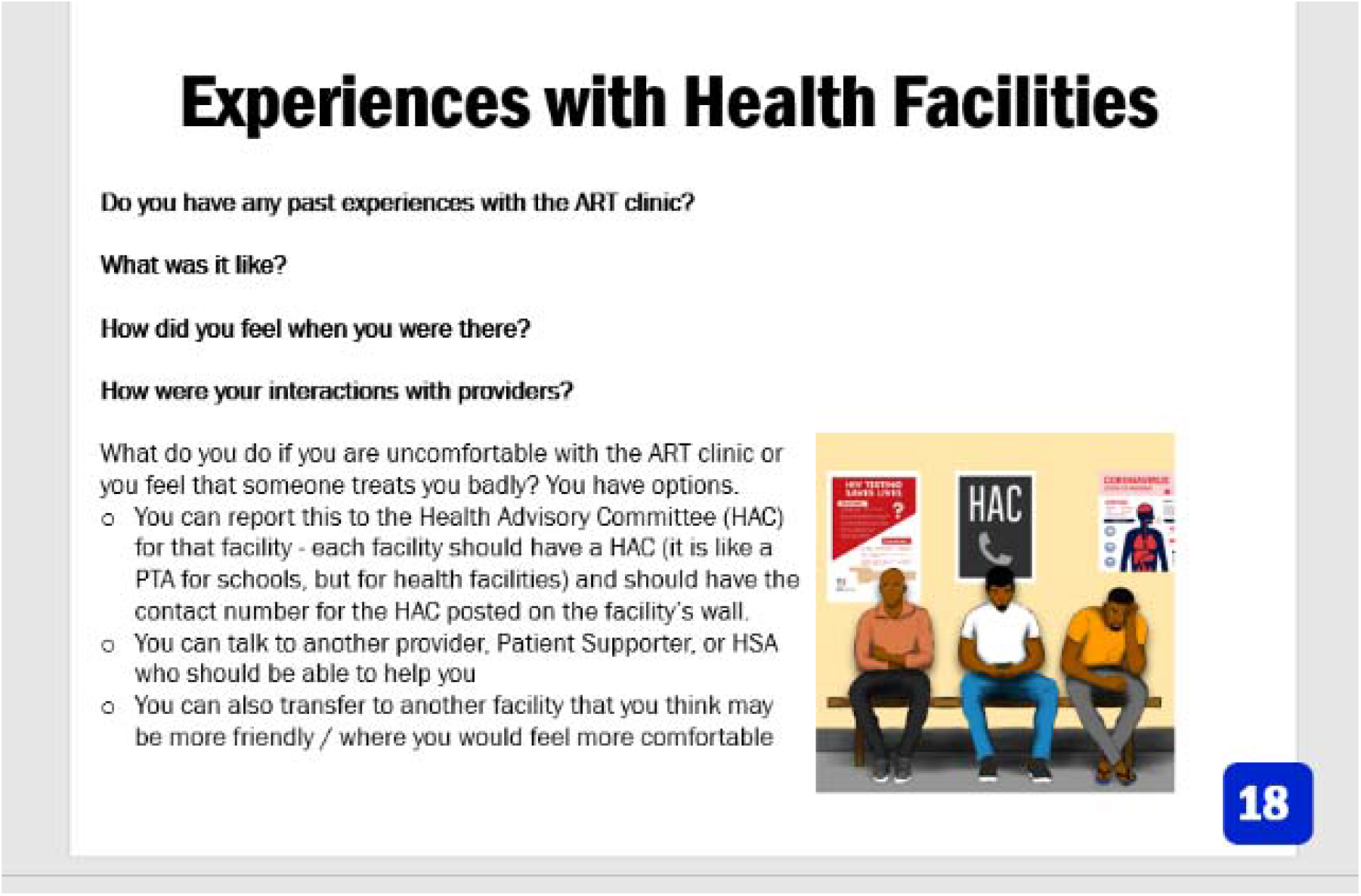
An example for the final product page

## Discussion

ART counselling curricula must be tailored to address specific barriers and build on facilitators relevant to sub-populations. We found that adapting standard ART counselling to men’s specific needs in Malawi was feasible and highly desired by men. We used a five-step process to develop ART counselling specifically for men in Malawi, using men’s own words and addressing men’s unique needs. Using a comprehensive approach, we identified the needs of men, gaps within standard counselling curriculum, and how to adapt standard curriculum. We aimed to meet men’s needs in ways that were accessible and motivating to men, being responsive to local context as reported by men we interviewed. We found that men needed to be treated as equals and engaged in planning and deciding on their health. However, they had limited knowledge on topics like how treatment contributes to their goals, navigating the health system, and self-compassion while on treatment. Standard counselling curriculum did not include these topics or person-centered counselling techniques essential to building trust between HCW and client [49–51] and self-efficacy within clients [52, 53]. Importantly, we found that men in Malawi had large gaps in knowledge regarding TaSP, benefits of dolutegravir and early ART initiation. Despite this, they were keen to know more about how to live well, given their local HIV epidemic. Men believed ART curriculum should incorporate local analogies and graphics representing local men in Malawi.

Counselling must promote and value individual autonomy and agency. In our study, men welcomed and accepted interactive, equal relationships with HCWs when they sought health services. In a previous study in Malawi[25], men expressed the need for counselling curricula that recognized them and allowed for their expression of thought and participation in health-related decisions. Men may experience even worse outcomes due to HCW bias against men as difficult clients[54, 55]. Some HCWs believe that men do not need extra help engaging in HIV services but actively choose not to access care as they are too stubborn and powerful[55, 56]. Such perceptions can reduce HCW motivation to actively engage men in counselling. A less didactic design and flexibility, and allowing clients to ask questions and debate information, would support the development of tailor-made sessions that respect men’s autonomy and allow them agency in decision making. This would improve men’s uptake of ART services and their adherence on ART.

We found that the men had major gaps in their knowledge about ART and the health system [5, 7]. Men were largely unaware about TaSP, its benefits and how it can contribute to their goals of a healthy family. TasP is a fairly new approach and current MoH counselling curriculum has not yet been updated to include the concept. This curriculum, with its strongly biomedical approach, fails to articulate both TasP and the benefits of early treatment in a manner that appeals to men’s needs. Including TasP messaging in the counselling curriculum not only addresses the knowledge gap, it acts as a motivator, resonating with men’s priorities to provide and protect their family[7, 36]. Facility navigation was another barrier to men’s engagement with health facilities[25]. The MoH curriculum did not include discussions on how men may deal with health system barriers including unsatisfactory care from providers. Including messaging about navigating health facilities in counselling curricula would familiarize men with the health system, thereby facilitating early treatment and adherence and addressing a health system barrier to care[7, 57].

Self-compassion has to be central in the counselling curriculum for men. Previous studies have identified lack of emotional readiness as a key barrier to men’s initiation and retention in ART care [11]. This may compound other challenges like stigma and fear of losing relationships, which also affect adherence [58]. In the context of ART, it has been argued that emotional readiness - an individual’s identification that treatment is beneficial - is an important predictor of adherence and retention[59]. The current MoH curriculum does not explore individual readiness, nor does it promote self-compassion to address individual-level barriers. A curriculum for male counselling must therefore promote individuals’ emotional readiness, status acceptance and self-compassion in their life-long treatment journey.

In order to be client-centered, counselling curriculum must also include pictures, prompts and job aids[60–62]. A recent review of medication argued for the use of images instead of didactic methods that depend on verbal and oral communication with patients[63]. The lack of counselling approaches that are client-centered affects everyone. It reduces the impact of counselling for both male and female clients [64]. Additionally, it can undermine HCW job satisfaction as there are few positive, meaningful interactions and reciprocity with their clients [65, 66]. With a biomedical and didactic approach, the current MoH counselling fails to provide client-centered counselling. It offers generic educational information and has less flexibility to adapt care interventions that utilize men’s motivators, meet their needs and address their barriers.

Our study demonstrates the feasibility of modifying a national counselling curriculum for the needs of a specific population. A limitation is that we focused broadly on men, but did not consider sub-populations of men, for example by age, sexual orientation or levels of absolute poverty. These factors may impact on health-seeking behaviors and attitudes towards HIV services [26, 35, 41], and certain populations may require further tailored counselling curriculum. Incorporating client-centered counselling [67, 68] and motivational interviewing techniques can help address this limitation. However, HCWs may still need to be trained on unique strategies to successfully engage sub-populations of men.

In conclusion, we found that it was possible to adapt a curriculum to suit men’s needs, thus addressing barriers to ART and support treatment initiation and adherence. Adapting a curriculum requires in-depth understanding of target users, and piloting the curriculum with target population.

## Data Availability

All data produced in the present work are contained in the manuscript

## Declarations

The IDEaL trial is supported by the Bill and Melinda Gates Foundation (INV-001423), The ENGAGE trial is supported by the National Institute for Mental Health (R01-MH122308) and the National Institute of Health Fogarty International Center (K01-TW011484-01). KD is supported by Health Fogarty International Center. We are very grateful for the contributions of all participating HCWs and study participants

## Compliance with Ethical Standards

The study protocol is approved by the institutional review boards of the University of California, Los Angeles (UCLA) (#21–000592) and the National Health Sciences Research Council (NHSRC) (#2562) in Malawi. The trial is registered with ClinicalTrials.gov as NCT04858243. There are no potential conflicts of interest to declare

